# Persistent racial/ethnic associated disparity in anti-tumor effectiveness of immune checkpoint inhibitors despite equal access

**DOI:** 10.1101/2021.11.26.21266821

**Authors:** M.A. Florez, J.O. Kemnade, N. Chen, W. Du, A. L. Sabichi, D.Y. Wang, Q. Huang, C.N. Miller-Chism, A Jotwani, A.C. Chen, D. Hernandez, V.C. Sandulache

## Abstract

Immune checkpoint inhibitors (ICIs) have revolutionized the treatment of both lung cancer and head and neck squamous cell carcinoma demonstrating clear benefit over traditional chemotherapy alone in the metastatic setting. While the overwhelming majority of ICI trial participants have been White patients, results of these trials have been broadly applied to patients of all ethnic/racial backgrounds. It has, therefore, not been well defined if response to ICIs differs between ethnic/racial populations or socio-economic groups.

We reviewed response to ICI of 208 patients with diagnoses of lung or head and neck cancers treated with ICI between 2015 and 2020 at one of three clinical pavilions associated with the Dan L. Duncan Comprehensive Cancer Center at Baylor College of Medicine in Houston, TX. Two of these pavilions (Harris Health System and the Michael E. DeBakey Veterans Affairs Medical Center) serve large minority patient populations and provide equal access of care to patients regardless of means. Of the 208 patients, 175 had a diagnosis of lung cancer [non-small cell lung carcinoma (NSCLC) or small cell lung cancer (SCLC)] and 33 had a diagnosis of head and neck squamous cell carcinoma (HNSCC); 38% self-identified as Black, 45% as non-Hispanic White, and 18% as Hispanic. The objective response rate (ORR) was similar for lung cancer (31.4%) and HNSCC patients (27.3%) (p=0.894). Statistically, the ORR for Hispanic and Black patients did not differ compared to non-Hispanic White patients (H 23.7%, B 28.6%, W 35.5%; H vs. W p=0.189; B vs. W p=0.338). When considering patients treated with ICI monotherapy, the ORR for Hispanic patients dropped to 13.3% and was significantly lower than the ORR of the non-Hispanic White patients while the ORR of Black and non-Hispanic White patients remained about the same (B 29.3% and W 34.6%, H vs. W p=0.0285; B vs. W p=0.5131). Immune related adverse events (irAEs) were the lowest in the Hispanic population occurring in only 30% of patients compared to 50% of patients exhibiting irAEs in the Black and non-Hispanic white cohorts.

ICIs demonstrate comparable anti-tumor effects in lung cancer (NSCLC + SCLC) and HNSCC during routine clinical practice regardless of race or ethnicity. The significantly lower ORR observed in our cohort for Hispanic patients, particularly when used as monotherapy, is an unexpected finding and will require additional study to identify potential biological and non-biological confounders which could contribute to reduced ICI effectiveness in this patient population.

## Introduction

Since the approval of the first immune checkpoint inhibitor (ICI) to treat melanoma in 2011^1^, ICIs targeting the programmed death (PD)-1 – programmed death ligand (PDL)-1 signaling axis between tumor cells and infiltrating T-lymphocytes have demonstrated activity in multiple solid tumor types.^2,3,4,5^ ICIs targeting PD-1 (e.g., nivolumab and pembrolizumab) and PD-L1 (e.g., atezolizumab, durvalumab) have been shown to improve survival in non-small cell lung cancer (NSCLC) patients, and have been established as part of standard-of-care therapy since 2015.^5-14^ Nivolumab and pembrolizumab were approved to treat head and neck squamous cell carcinoma (HNSCC), another smoking related malignancy, in 2016 and 2018, respectively.^15,16^

Although the potential of ICIs to improve disease control and survival is now well established, whether the benefit of ICIs can be realized at a population level remains an open question given persistent disparities in cancer diagnosis and treatment delivery for patients with advanced stage and metastatic disease.^17-22^ Unfortunately, despite persistent efforts, <25% of cancer-related clinical trial participants are racial minorities.^23-25,26^ This inclusion disparity is particularly obvious in clinical trials that led to the approval of ICIs for the treatment of NSCLC (Checkmate 017, 90% White^7^ ; Checkmate 057, 91% White^6^ ; Keynote 001, 82% White^9^; OAK, 71% White^13^; Impower 150, 82% White^14^; Keynote 010, 73% White^10^) and in HNSCC (Checkmate 141, 83% White^27^; Keynote 048, 73% White^16^). Compounding this deficit is the fact that most cancer-related phase III clinical trials fail to report ethnicity.^23^

Given the limited data available regarding ICI effectiveness in minority patients, some investigators have attempted to address this issue by comparing the safety and efficacy of ICI treatment for NSCLC in real world populations. Ayers *et al*. found a trending survival benefit in African Americans treated with ICI in a cohort consisting of 30% African Americans (HR 0.6; P=0.062)^25^, while Nazha *et al*. reported no difference in overall survival with ICI treatment in African Americans (P=0.84).^28^ Data on Hispanic patients, now the largest minority population in the United States is lacking from all studies to date, whether prospective or retrospective.^29^ The Harris Health System (HHS) and the Michael E. DeBakey Veterans Affairs Medical Center (MEDVAMC) in Houston, Texas provide tertiary cancer care for a diverse patient population (e.g. HHS-57% Hispanic, 25% African American, 10% White, and 8% other) providing equal access to Harris County residents or Veterans, respectively, regardless of insurance status or financial means. We retrospectively analyzed oncologic outcomes for NSCLC and HNSCC patients treated with PD-1 and PDL-1 ICIs at these equal access institutions in order to determine whether racial and/or ethnic disparities are present.

## Methods

### Patient Population

Following approval from Baylor College of Medicine (BCM), HHS and MEDVAMC Institutional Review Boards, we performed a retrospective analysis of patients with a diagnosis of NSCLC or HNSCC between 2015 and 2020. Patients had either locally recurrent disease for which definitive local therapy (surgery or radiation) was no longer an option or metastatic disease. Patients receiving adjuvant immunotherapy as part of a definitive treatment strategy were excluded from the analysis. All collection and analysis of the current data was performed in a manner consistent with existing standards for clinical research (Declaration of Helsinki, US Federal Policy for the Protection of Human Subjects). Patient race/ethnicity (self-identified and listed in the electronic medical record), age, gender, tobacco and alcohol exposure, tumor characteristics, treatment history, ICI toxicities, response to ICI, PDL-1 status, ECOG performance status, and BMI were collected and analyzed. Tumor histology, stage (T, N, M-classifications), number of metastases, radiation treatment, and chemotherapy treatment were collected through review of the electronic medical records. Inclusion criteria included: 1) primary NCSLC or HNSCC, 2) tissue diagnosis at the participating institutions, 3) treatment delivery at the participating institutions, 4) treatment with ICI or ICI plus conventional cytotoxic chemotherapy. Patient response to ICI was determined by review of surveillance imaging (PET/CT, CT, or MRI; using official radiologist/nuclear medicine reading) following ICI treatment to determine complete response, partial response, stable disease, or progressive disease. Routine images were reviewed for each patient up to death or loss to follow up (LTFU) to determine best overall response. Best overall objective response rate (ORR) was determined by calculating the number of patients attaining a complete response or a partial response and dividing by the total patient population. PD-L1 status was ascertained from clinical records using companion testing for ICIs for those lung cancer patients for which data was available and reported as percentage of viable tumor cells expressing the protein.

### Statistical analysis

In order to assess the efficacy of ICIs with as few patient and disease intrinsic confounders as possible, we chose ORR as the primary endpoint for comparison. Associations between identified clinical, biological and pathologic variables were determined by two-sided Chi-square tests. Statistical calculations were performed with Prism (Graphpad Software LLC Version 9.1.2). For all statistics, p-values were considered to be statistically significant if below a threshold of 0.05 (two-sided).

## Results

### Patient and disease characteristics

We analyzed data for 175 lung and 33 head and neck cancer patients receiving either ICI therapy alone or in combination with chemotherapy across all three treatment pavilions at BCM. As the goal of our study was to compare treatment responses of the Black and Hispanic patient population to those of the White patient population, **Table 1** details the patient demographics and disease characteristics categorized by these three different racial/ethnic cohorts (**Table 1, Supplementary Table 1**). A slight majority of the patients were non-Hispanic White (45%) followed closely by Black patients (37%) while the Hispanic cohort constituted 18% of the study population. The three cohorts were generally evenly matched among the detailed patient and disease characteristics with significant differences found in three categories among the entire cohort and two categories among the ICI monotherapy cohort. Specifically, the Hispanic population consisted of fewer smokers compared to the White population (p=0.0003) and had more sites of metastatic disease compare to the White patients (p=0.0224). The White population reported more alcohol consumption than Black patients (p=0.0272) (**Supplementary Figure 1A-C**). These differences were also present in the cohort of patients that received ICI monotherapy, except for alcohol intake, which was similar between the various racial/ethnic cohorts (**Supplementary Figure 2A & B)**.

**Table 1.** Patient and treatment characteristics.

|  |  | **Black** | **White** | **Hispanic** |
| --- | --- | --- | --- | --- |
| **N** |  | 77 | 93 | 38 |
| **Age at diagnosis** |  | 61.4 | 62.4 | 59.6 |
| **Age at ICI initiation** |  | 62.8 | 64.3 | 60.6 |
| **Sex** |  |  |  |  |
|  | Male | 58 (75.3) | 69 (74.2) | 23 (60.5) |
|  | Female | 19 (24.7) | 24 (25.8) | 15 (39.5) |
| **Smoking status** |  |  |  |  |
|  | Yes | 66 (85.7) | 85 (91.4) | 25 (65.8) |
|  | Never smoker | 10 (13.0) | 8 (8.6) | 13 (34.2) |
|  | Smoker at diagnosis | 21 (27.3) | 34 (36.6) | 7 (18.4) |
|  | Unknown | 1 (1.3) | 0 (0) | 0 (0) |
| **Alcohol** |  |  |  |  |
|  | Yes | 21 (27.3) | 39 (41.9) | 11 (28.9) |
|  | No | 50 (64.9) | 44 (47.3) | 26 (68.4) |
|  | Unknown | 6 (7.8) | 11 (11.8) | 0 (0) |
| **Cancer** |  |  |  |  |
|  | Lung | 68 (88.3) | 75 (80.6) | 32 (84.2) |
|  | Adenocarcinoma | 51 (75.0) | 44 (58.7) | 23 (71.9) |
|  | Squamous | 15 (22.1) | 21 (28.0) | 7 (21.9) |
|  | Small cell | 2 (2.9) | 8 (10.7) | 2 (6.3) |
|  | NSCLC (NOS) | 0 (0) | 2 (2.7) | 0 (0) |
|  | Head and neck SCC | 9 (11.7) | 18 (19.4) | 6 (15.8) |
| **ECOG** |  |  |  |  |
|  | 0 | 9 (11.7) | 13 (14.0) | 8 (21.1) |
|  | 1 | 33 (42.9) | 40 (43.0) | 19 (50.0) |
|  | 2 | 23 (29.9) | 27 (29.0) | 7 (18.4) |
|  | 3 | 3 (3.9) | 7 (7.5) | 3 (7.9) |
|  | 4 | 1 (1.3) | 0 (0) | 0 (0) |
|  | Unknown | 8 (10.4) | 6 (6.5) | 1 (2.6) |
| **Number of Metastasis** |  |  |  |  |
|  | 0 | 10 (13.0) | 12 (12.9) | 5 (13.2) |
|  | Contralateral lung only | 7 (9.0) | 4 (4.3) | 6 (15.8) |
|  | 1 | 20 (26.0) | 41 (44.1) | 4 (10.5) |
|  | 2 | 30 (39.0) | 16 (17.2) | 15 (39.5) |
|  | 3 | 6 (7.8) | 16 (17.2) | 5 (13.2) |
|  | 4+ | 4 (5.2) | 4 (4.3) | 3 (7.9) |
|  | Any brain metastasis | 13 (16.9) | 25 (26.9) | 13 (34.2) |
| **Prior systemic therapy (all)** |  |  |  |  |
|  | 0 | 19 (24.7) | 24 (25.8) | 10 (26.3) |
|  | 1 | 33 (42.9) | 46 (49.5) | 18 (37.4) |
|  | 2 | 18 (23.4) | 15 (16.1) | 5 (13.2) |
|  | 3 | 4 (5.2) | 4 (4.3) | 2 (5.2) |
|  | 4+ | 3 (3.9) | 4 (4.3) | 3 (7.9) |
| **Prior systemic therapy (concurrent and adjuvant chemo excluded)** |  |  |  |  |
|  | 0 | 29 (37.7) | 39 (41.9) | 17 (44.7) |
|  | 1 | 33 (42.9) | 41 (44.1) | 14 (36.8) |
|  | 2 | 10 (13.0) | 6 (6.5) | 2 (5.3) |
|  | 3 | 2 (2.6) | 3 (3.2) | 2 (5.3) |
|  | 4+ | 2 (2.6) | 4 (4.3) | 3 (7.9) |
| **Concurrent chemo with ICI** |  |  |  |  |
|  | Yes | 19 (24.7) | 15 (16.1) | 8 (21.1) |
|  | No | 58 (75.3) | 78 (83.9) | 30 (78.9) |
| **Prior radiation anywhere** |  |  |  |  |
|  | Yes | 49 (63.6) | 73 (78.5) | 22 (57.9) |
|  | No | 28 (36.4) | 20 (21.5) | 16 (42.1) |
| **PD-L1 status (NSCLC only)** |  |  |  |  |
|  | 0% | 7 (10.6) | 8 (11.9) | 5 (16.7) |
|  | 1-49% | 17 (25.8) | 8 (11.9) | 6 (20.0) |
|  | >50% | 16 (24.2) | 18 (26.9) | 10 (33.3) |
|  | Unknown | 26 (39.4) | 41 (61.2) | 11 (36.7) |

### Treatment characteristics

Treatment characteristics were well matched among the racial/ethnic cohorts **(Table 1)**. About 20% of patients among each cohort received ICI in combination with chemotherapy. 40% of the patients in each cohort received ICI as first line treatment for recurrent or metastatic disease. Both Hispanic and Black patients were subject to less radiation treatment (definitive or palliative) than White patients (p=0.0165 and p=0.0322, respectively) (**Supplementary Figure 1D)**.

The ICI monotherapy patient population also showed similar treatment characteristics among the three racial/ethnic groups (**Supplementary Table 1)**. Only 20% of these patients received ICI monotherapy in the first-line setting. Also, only the Hispanic population was subject to significantly less radiation therapy (definitive or palliative) than the White population in this treatment cohort (p=0.026) (**Supplementary Figure 2C)**.

### Overall population responses

Treatment responses were first calculated for the entire cohort and compared by disease type. ORR was approximately 30% for the entire 208 patient cohort and was similar between the NSCLC and HNSCC patient cohorts (**Figure 1a**). Overall, the response rates were similar between the cohorts that received ICI monotherapy and ICI in combination with chemotherapy (**Figure 1b**). PD-L1 status was available for slightly more than half of NSCLC patients reviewed (**Table 1**). As expected, a PD-L1 status of 50% or more was associated with a significantly (p=0.0029) higher ORR compared to a PD-L1 less than 50% in the population of patients receiving ICI monotherapy (**Figure 1c**). Given some significant differences in patient, disease, and treatment characteristics between the racial cohorts noted above, we calculated ORRs for each of these differences separately to assure they did not confound the ORR analysis of the racial cohorts detailed below. ORRs did not differ significantly for any of the disparate characteristics described in either the entire study cohort or the ICI monotherapy cohort (**Supplementary Figures 3 and 4)**.

**Figure 1.**
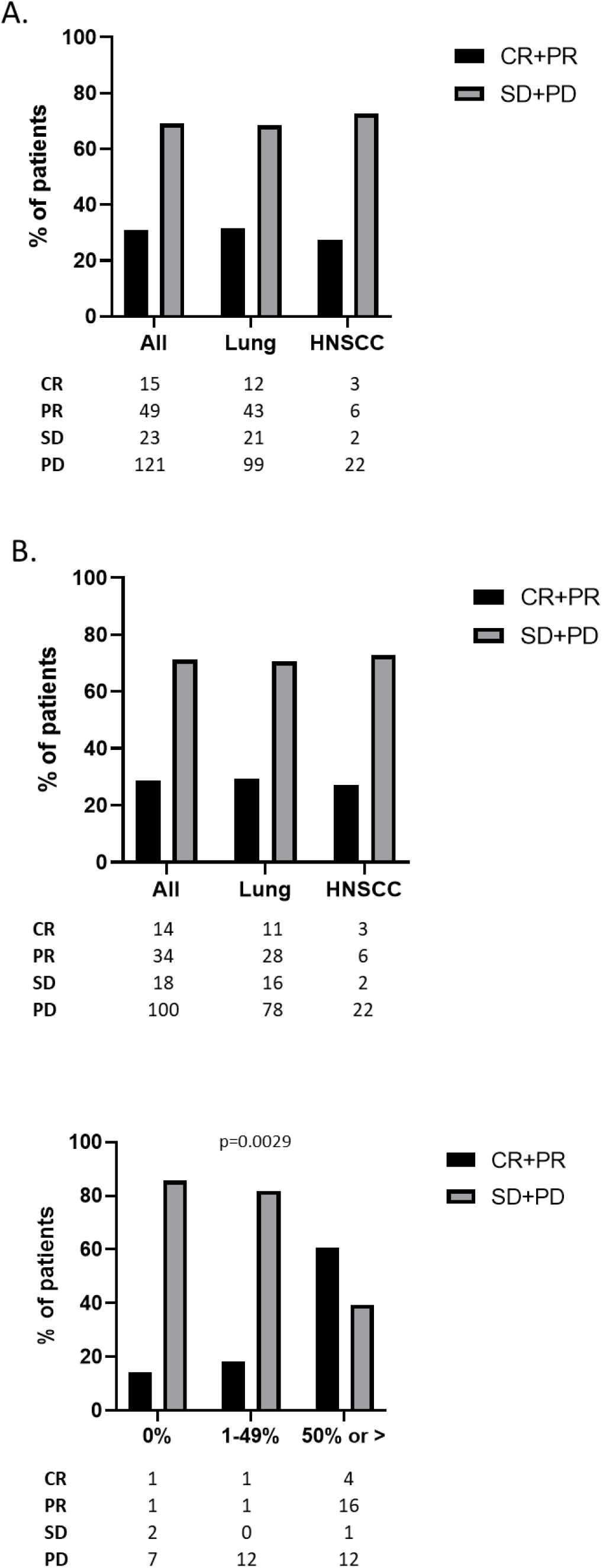
Best overall response. **A)** ORR data for all patients and by disease category for patients treated with either ICI monotherapy or chemo/ICI combination therapy. **B)** ORR data for all patients and by disease category for patients treated with ICI monotherapy. **C)** ORR data for NSCLC patients stratified by PD-L1 status. Only patients for whom PD-L1 was known were included. CR- complete response; PR- partial response; SD- stable disease; PD- progressive disease.

### Responses by race/ethnicity

While the ORR in Hispanic patients was lower than that of Black and non-Hispanic White patients (23.7% compared to 28.6% and 35.5%, respectively) these differences were not statistically significant compared to the non-Hispanic White patient population (**Figure 2a**). However, when patients receiving chemo and ICI combination therapy were removed from the analysis, the ORR of the Hispanic cohort decreased significantly. Only 13.3% of Hispanic patients treated with ICI monotherapy had a response while the ORR for Black and non-Hispanic White patients remained about the same, 29.3% and 34.6% respectively. Compared to non-Hispanic White patients, the ORR of the Hispanic patient cohort was significantly less while the ORR of the Black patient cohort did not differ significantly (**Figure 2b**). More than three-fourths of Hispanic patients’ tumors were refractory to ICI monotherapy while lack of response to ICI monotherapy was seen in only 55.2% and 57.7% of Black and non-Hispanic White patients, respectively. No complete responses (CR) were observed in the Hispanic ICI monotherapy cohort and only one patient treated with combination immunotherapy and chemotherapy achieved a CR. Black and non-Hispanic White patients treated with ICI monotherapy had a CR rate of 5.7% and 14.1%, respectively (**Figure 2b**). Interestingly, all the CRs in the Black and non-Hispanic White populations were in patients treated with ICI monotherapy.

**Figure 2.**
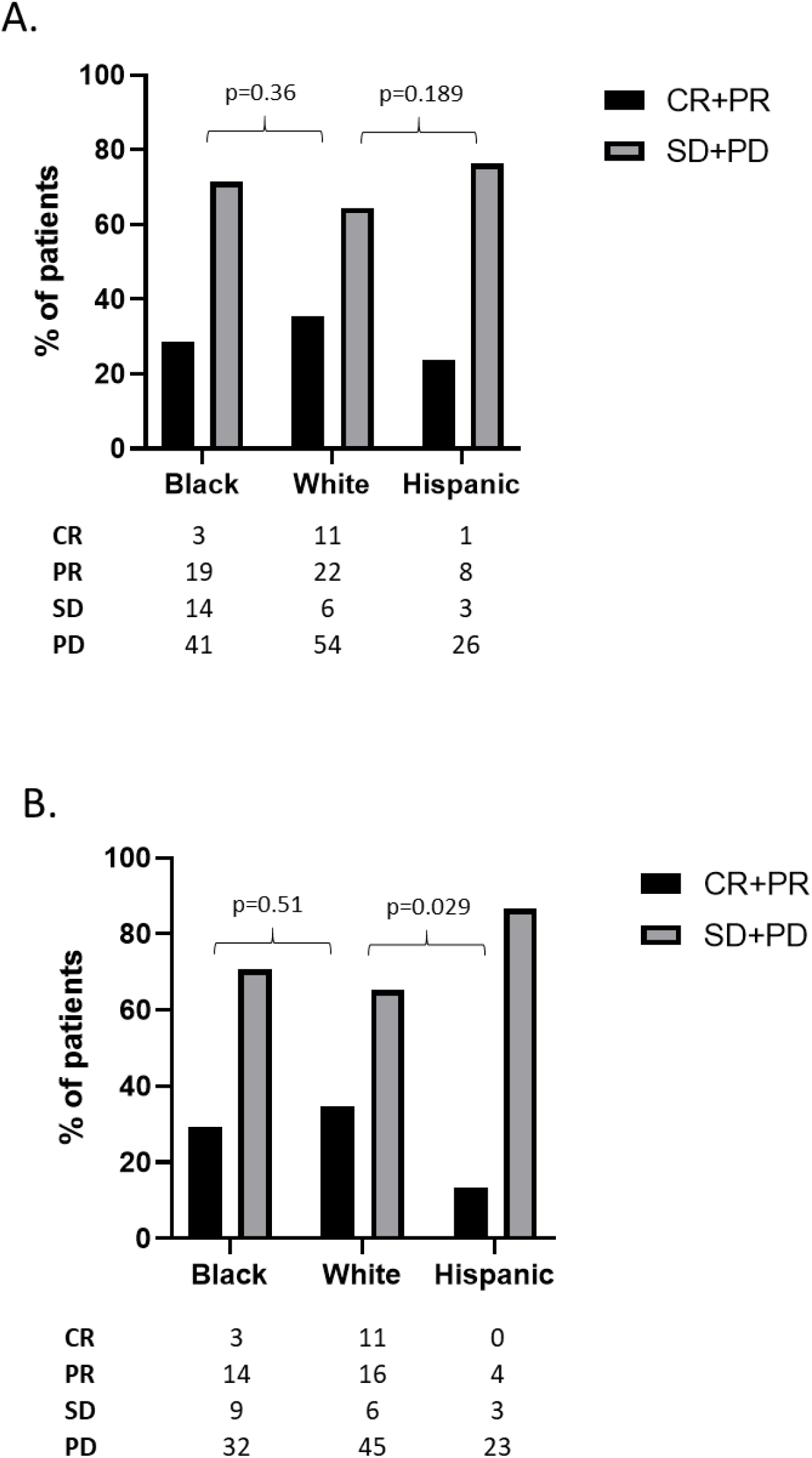
Best overall response by race/ethnic group. **A)** ORR data for all patients and by race/ethnic group for patients treated with either ICI monotherapy or chemo/ICI combination therapy. **B)** ORR data for all patients and by race/ethnic group for patients treated with ICI monotherapy. CR- complete response; PR- partial response; SD- stable disease; PD- progressive disease.

### Responses by PD-L1 in NSCLC

We assessed ORR in NSCLC patients using a combination of PD-L1 status and race/ethnicity. Again, as expected, Black and non-Hispanic White patients with a PD-L1 of 50% or greater exhibited a high ORR of over 60% when treated with ICI monotherapy (**Figure 3a**). This ORR dropped drastically to approximately 20% or below for tumors with PD-L1 less than 50% (**Figures 3b, c**). In our analysis, the ORR of NSCLC in Hispanic patients treated with ICI monotherapy was less than 30% regardless of PD-L1 expression (**Figure 3**).

**Figure 3.**
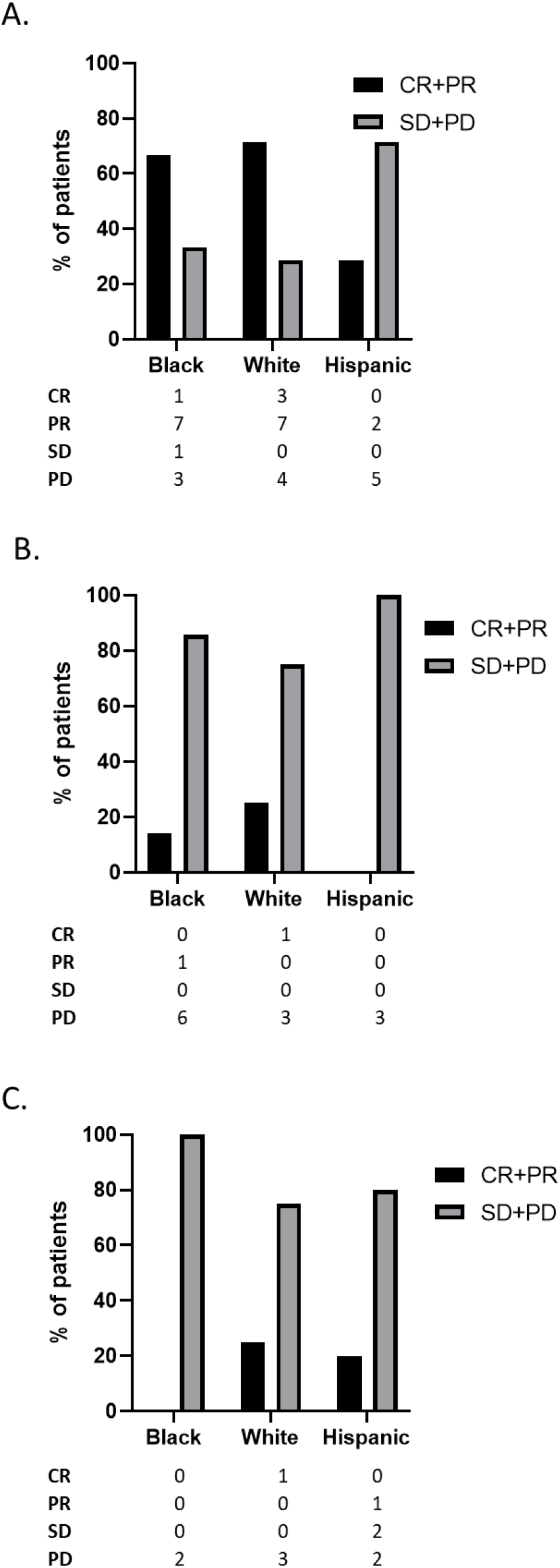
Best overall response by PD-L1. ORR data by race/ethnic group for NSCLC patients treated with ICI monotherapy stratified by PD-L1 **A**) ≥50%, **B**) 1-49% and **C**) 0%. CR- complete response; PR- partial response; SD- stable disease; PD- progressive disease.

### Immune-related Adverse Events

Adverse events thought to be related to immunotherapy (irAEs) were analyzed for the ICI monotherapy cohort. Approximately 50% of Black and non-Hispanic White patients experienced irAEs of any grade. However, only 30% of Hispanic patients developed any irAEs which was significantly less (p=0.0121) than non-Hispanic White patients (**Figure 4a**). We assessed the frequency of severe irAEs defined as toxicities resulting in systemic steroid administration, temporary cessation of ICI therapy, or permanent cessation of therapy among the three racial/ethnic cohorts. Approximately 15-20% of patients, regardless of race/ethnicity, experienced severe irAEs with about 10% of patients, again regardless of race/ethnicity, requiring a complete cessation of therapy (**Figure 4b**). Given the shared mechanism of ICI irAEs and anti-tumor immunity, we hypothesized that the presence of toxicity would predict positive response to ICIs. Indeed, patients experiencing at least one irAE in the ICI monotherapy cohort had a significantly higher ORR than those patients exhibiting no irAEs (**Figure 4c**) (p=0.049).

**Figure 4.**
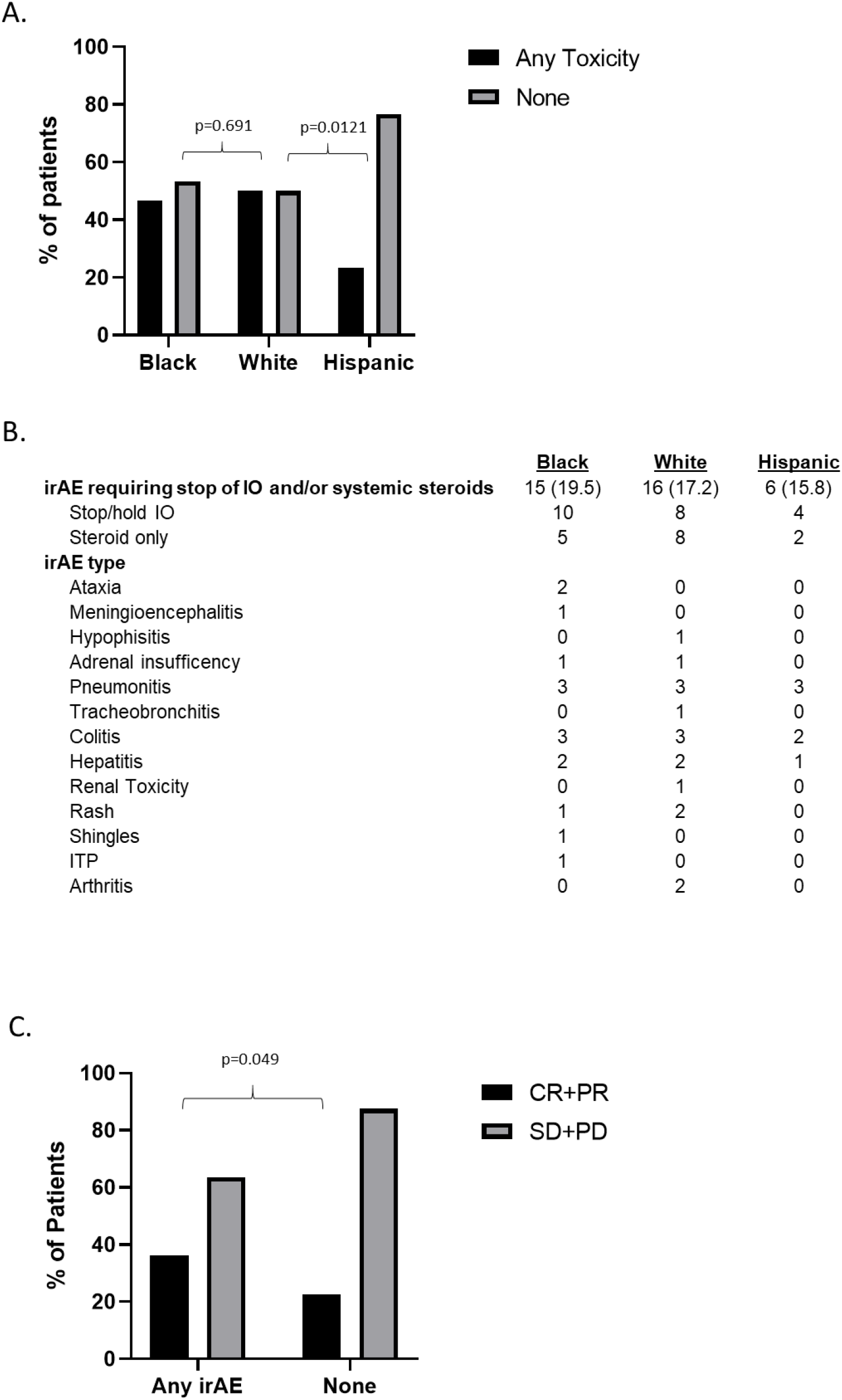
Immune-related adverse events (irAEs) within the ICI monotherapy cohort. **A)** Overall irAE rate as a function of race/ethnic group. **B)** Severe irAEs resulting in steroid administration or ICI cessation with summary of specific severe irAEs. **C)** ORR as a function of irAE for the ICI monotherapy cohort. CR: Complete response; PR: Partial response; SD: stable disease; PD: Progressive disease.

## Discussion

Although racial disparities persist across multiple solid tumor histologies in the US population at large, individual institutional studies have been able to demonstrate equivalence in the setting of equal access. Our previous work in HNSCC has shown that both HHS and the MEDVAMC are able to generate equivalent oncologic outcomes for patients which is consistent with data from other institutions.^30-34^ These data would suggest that cancer outcome disparities at a population level are likely driven primarily by socioeconomic status, unequal access to timely and high-quality healthcare, and other social factors. Indeed, retrospective studies have shown that race strongly correlates to HNSCC disease stage at presentation, with Blacks presenting more often with advanced locoregional disease or with metastatic disease.^19^ Similarly, for HNSCC some have suggested that reduced survival in Black patients is a function of differential distribution of low-risk and high-risk disease (as a function of human papillomavirus (HPV)) status.^18^ However, contradicting these assertions are data which indicate that even when HPV status is accounted for, Black HNSCC patients and to a lesser degree Hispanic patients demonstrate reduced survival compared to their counterparts.^35^ Other biological factors have also been considered. Ramakodi *et al*. identified ancestry related single nucleotide polymorphisms (SNPs) in the DNA polymerase beta gene which may impact response to conventional chemo-radiation strategies used for both HNSCC and NCSLC in the curative intent setting.^36^ Differential SNP distribution may be compounded by differential somatic mutation distribution in some HNSCC patients to further impede treatment response and reduce survival.^37^ Transcriptional data from African American patients demonstrates race-related shifts in both tumor metabolism and DNA repair across multiple disease sites including HNSCC and other smoking related malignancies.^38^

Taken together these data highlight an outstanding question for clinicians and patients alike: are Black and Hispanic patients likely to derive the same benefit from ICIs once access is established? Certainly, the sparsity of minority patients in prospective clinical trials and the lack of consistently reported race-based response data prevents a definitive answer using level I data. It is therefore critical that continued attempts to answer this question be made in the real-world, post-approval setting. This study offers retrospective data from our academic institution experience that represents the more diverse patient population that may be encountered in an urban center. Overall, our data are encouraging for Black patients. Not only are response rates comparable with those of White patients, but overall toxicity appears to be similar. Despite reports that show ICI enhances OS in Hispanics when compared to chemotherapy,^39^ our findings are concerning, particularly given the low rate of complete response. This lower response rate paralleled by a lower rate of irAEs suggest the potential for reduced effectiveness of this agent class. This report is the first showing a significant impact on ICI effectiveness in a substantial Hispanic NSCLC and HNSCC patient population and our findings will need to be validated in additional series. Specifically, to better understand this finding, future studies will need to be enriched for patients treated with ICI monotherapy. Furthermore, it will be essential to determine what other factors might contribute to the altered effectiveness of ICI in Hispanics. Studies have suggested that the efficacy of ICI in various cancer types may be impacted by multiple factors including the use of antibiotics,^40,41^ the gut microbiome, mutational burden, infections, and exercise.^42,43^ Thus, future studies will also be needed to assess the impact of additional factors on racial/ethnic disparities in ICI therapy.

We acknowledge the limitations of our study which include its retrospective nature, cohort size, limited availability of PD-L1 data for the NSCLC patients, and absence of detailed socio-economic/cultural cohort data. However, the cohort spans two distinct equal access institutions with significant minority populations, affiliated with a tertiary academic institution and an NCI-designated comprehensive cancer center (CCC), helping control for at least one important factor contributing to oncologic outcome disparities among racial minorities: access to care. To further evaluate the outcomes of this study, a concerted effort is needed to enact specific policies and funding opportunities focused on minority recruitment in order to encourage enrollment of minority patients into clinical trials focused on ICIs in NSCLC and HNSCC in a manner representative of current demographic shifts. As such, we consider it imperative that NCI-designated CCCs which serve predominantly minority populations form a closer network designed for data sharing, specimen banking and research integration. Within such a network, it is important to support biobanking and genotyping efforts that will allow us to develop biologically focused analyses of interactions between ancestry and treatment response. These analyses may help to elucidate the effects of socio-economic/cultural/environmental factors compared to biological differences which even in the context of equal access to care is not adequately parsed out. Furthermore, patient diversity in these efforts is especially important as major existing bio-response databases predominantly consist of White population of European ancestry.^44^ Combined with national efforts such as those exemplified by the Million Veteran Program which seeks to elucidate the health impacts of genetics, lifestyle, and military exposure, such a network could help better understand how to serve and advance the health of our patients In conclusion, this retrospective cohort study identifies a potential signal of decreased ICI-response in Hispanic lung cancer and HNSCC patients. Coupled with a significantly lower irAE rate for Hispanic patients, the data suggest a possible underlying mechanistic reason for this disparity. However, the exact causes remain unclear and may be linked to differences equally as disparate ranging from variations in intrinsic tumor biology and immunology, genetics and epigenetics to extrinsic socio-economic and cultural (e.g. diet, exercise, medication exposure, etc.) factors. Expanding the investigation of this outcome and its causes is an imperative next step in the effort to improve outcomes for Hispanic lung and head and neck cancer patients and will serve to broaden our understanding of factors impacting ICI response.

## Supporting information

EQUATOR Network research reporting checklist

## Data Availability

All data produced in the present work are contained in the manuscript

