## Supplemental Data for "Persistent racial/ethnic associated disparity in anti-tumor effectiveness of immune checkpoint inhibitors despite equal access"

### Supplementary Data

#### Supplementary table 1. Patient and treatment characteristics of ICI monotherapy cohort.

| | | Black | White | Hispanic |
|---|---|---|---|---|
| **N** | | 58 | 78 | 30 |
| **Age at diagnosis** | | 62.8 | 63.2 | 60.6 |
| **Age at ICI initiation** | | 64.3 | 65.4 | 61.7 |
| **Sex** | | | | |
| | Male | 47 (81.0) | 61 (78.2) | 18 (60.0) |
| | Female | 11 (19.0) | 17 (21.8) | 12 (40.0) |
| **Smoking status** | | | | |
| | Yes | 48 (82.8) | 70 (89.7) | 20 (66.7) |
| | Never smoker | 9 (15.5) | 8 (10.3) | 10 (33.3) |
| | Smoker at diagnosis | 14 (24.1) | 31 (39.7) | 7 (23.3) |
| | Unknown | 1 (1.7) | 0 (0) | 0 (0) |
| **Alcohol** | | | | |
| | Yes | 17 (29.3) | 35 (44.9) | 11 (36.7) |
| | No | 36 (62.1) | 33 (42.3) | 19 (63.3) |
| | Unknown | 5 (8.6) | 10 (12.8) | 0 (0) |
| **Cancer** | | | | |
| | Lung | 49 (84.5) | 60 (76.9) | 24 (80.0) |
| | Adenocarcinoma | 34 (69.4) | 36 (60.0) | 18 (75.0) |
| | Squamous | 13 (26.5) | 15 (25.0) | 6 (25.0) |
| | Small cell | 2 (4.1) | 6 (10.0) | 0 (0) |
| | NSCLC (NOS) | 0 (0) | 3 (5.0) | 0 (0) |
| | Head and neck SCC | 9 (18.4) | 18 (23.1) | 6 (20.0) |
| **ECOG** | | | | |
| | 0 | 7 (12.1) | 12 (15.4) | 7 (23.3) |
| | 1 | 22 (37.9) | 32 (41.0) | 14 (46.7) |
| | 2 | 17 (29.3) | 22 (28.2) | 5 (16.7) |
| | 3 | 3 (5.2) | 6 (7.7) | 3 (10.0) |
| | 4 | 1 (1.7) | 0 (0) | 0 (0) |
| | Unknown | 8 (13.8) | 6 (7.7) | 1 (3.3) |
| **Number of Metastasis** | | | | |
| | 0 | 8 (13.8) | 11 (14.1) | 4 (13.3) |
| | Contralateral lung only | 4 (6.9) | 2 (2.6) | 5 (16.7) |
| | 1 | 17 (29.3) | 37 (47.4) | 1 (3.3) |
| | 2 | 21 (36.2) | 14 (17.9) | 15 (50.0) |
| | 3 | 4 (6.9) | 11 (14.1) | 4 (13.3) |
| | 4+ | 4 (6.9) | 3 (3.8) | 1 (3.3) |
| | Any brain metastasis | 9 (15.5) | 18 (23.1) | 8 (26.7) |

| | | | | |
|---|---|---|---|---|
| **Prior systemic therapy (all)** | | | | |
| | 0 | 9 (15.5) | 14 (17.9) | 5 (16.7) |
| | 1 | 28 (48.3) | 42 (53.8) | 17 (56.7) |
| | 2 | 15 (25.9) | 14 (17.9) | 4 (13.3) |
| | 3 | 4 (6.9) | 4 (5.1) | 1 (3.3) |
| | 4+ | 2 (3.4) | 4 (5.1) | 3 (10.0) |
| **Prior systemic therapy (concurrent and adjuvant chemo excluded)** | | | | |
| | 0 | 12 (20.7) | 21 (26.9) | 6 (20.0) |
| | 1 | 29 (50.0) | 43 (55.1) | 16 (53.3) |
| | 2 | 12 (20.7) | 7 (9.0) | 4 (13.3) |
| | 3 | 3 (5.2) | 3 (3.8) | 1 (3.3) |
| | 4+ | 2 (3.4) | 4 (5.1) | 3 (10.0) |
| **Prior radiation anywhere*** | | | | |
| | Yes | 39 (67.2) | 63 (80.8) | 18 (60.0) |
| | No | 19 (32.8) | 15 (19.2) | 12 (40.0) |
| **PD-L1 status (NSCLC)** | | | | |
| | 0% | 2 (4.3) | 4 (7.4) | 5 (20.8) |
| | 1-49% | 7 (14.9) | 4 (7.4) | 3 (12.5) |
| | >50% | 12 (25.5) | 14 (25.9) | 7 (29.2) |
| | Unknown | 26 (55.3) | 32 (59.3) | 9 (37.5) |

**Supplementary Figure 1. Cohort characteristics with significant differences among the race/ethnic groups for the entire cohort (ICI monotherapy and chemo/ICI combination therapy). A)** Smoking status, **B)** alcohol consumption, C**)** number of metastatic sites, and **D)** history of radiation treatment as a function race/ethnicity.

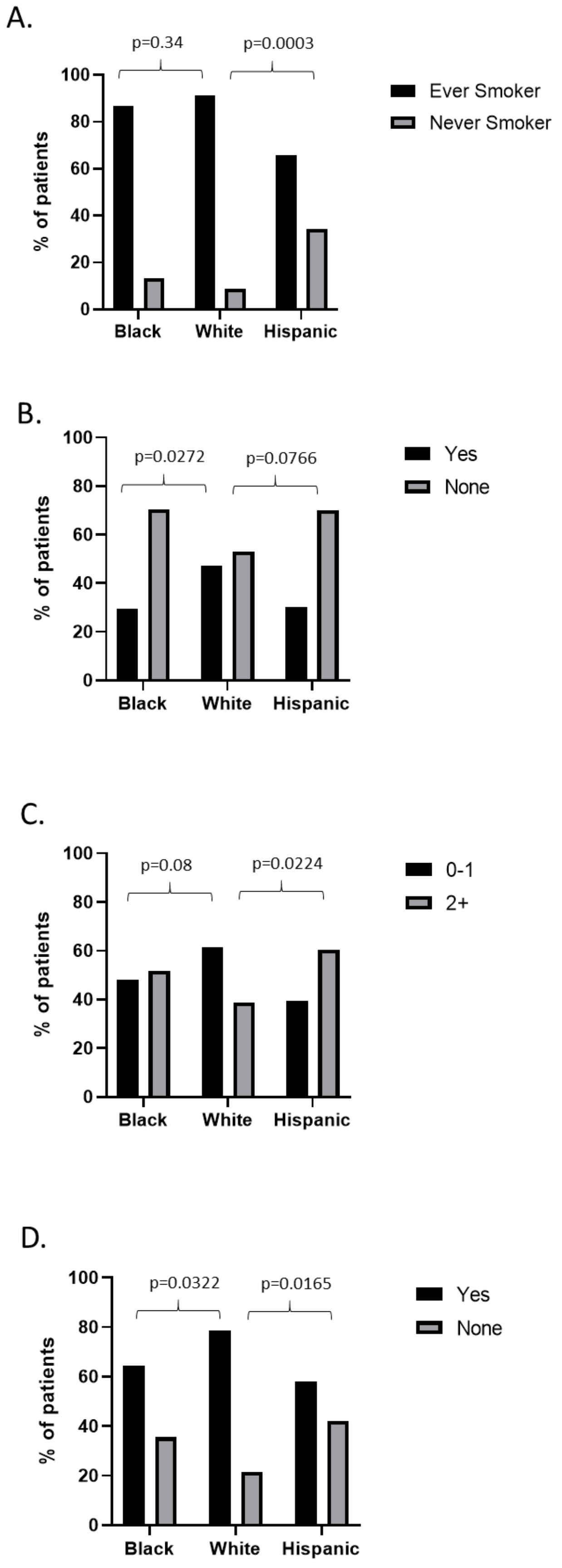

**Supplementary Figure 2. Cohort characteristics with significant differences among the race/ethnic groups for the ICI monotherapy cohort. A)** Smoking status, **B)** number of metastatic sites, and **C)** history of radiation treatment as a function race/ethnicity.

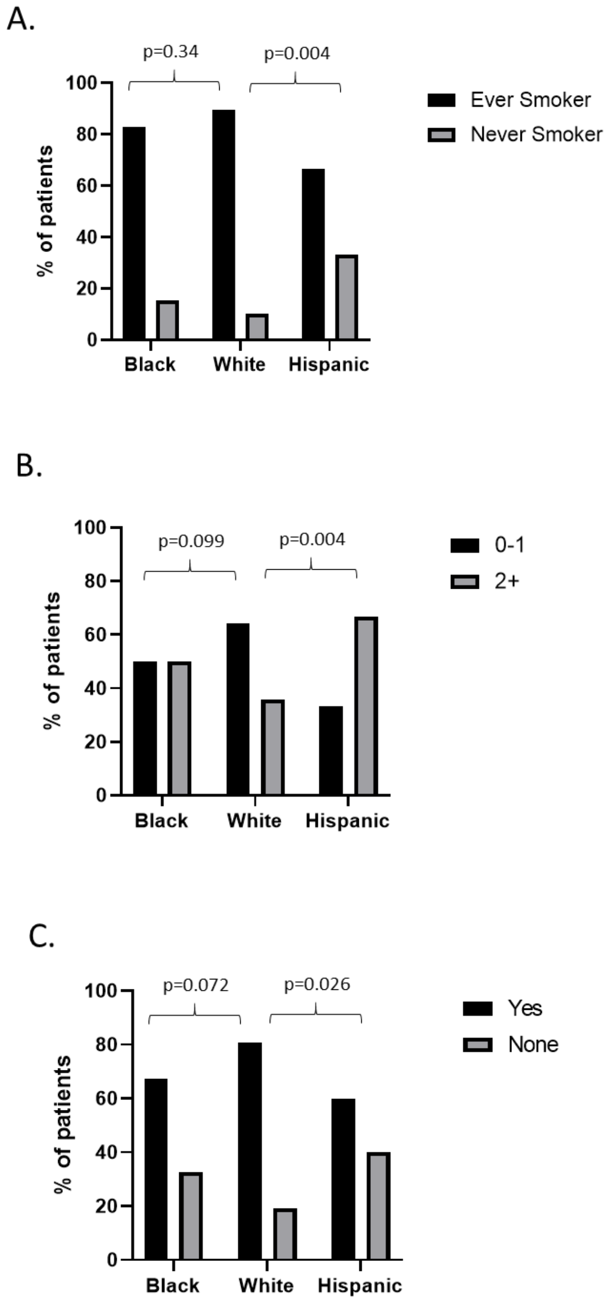

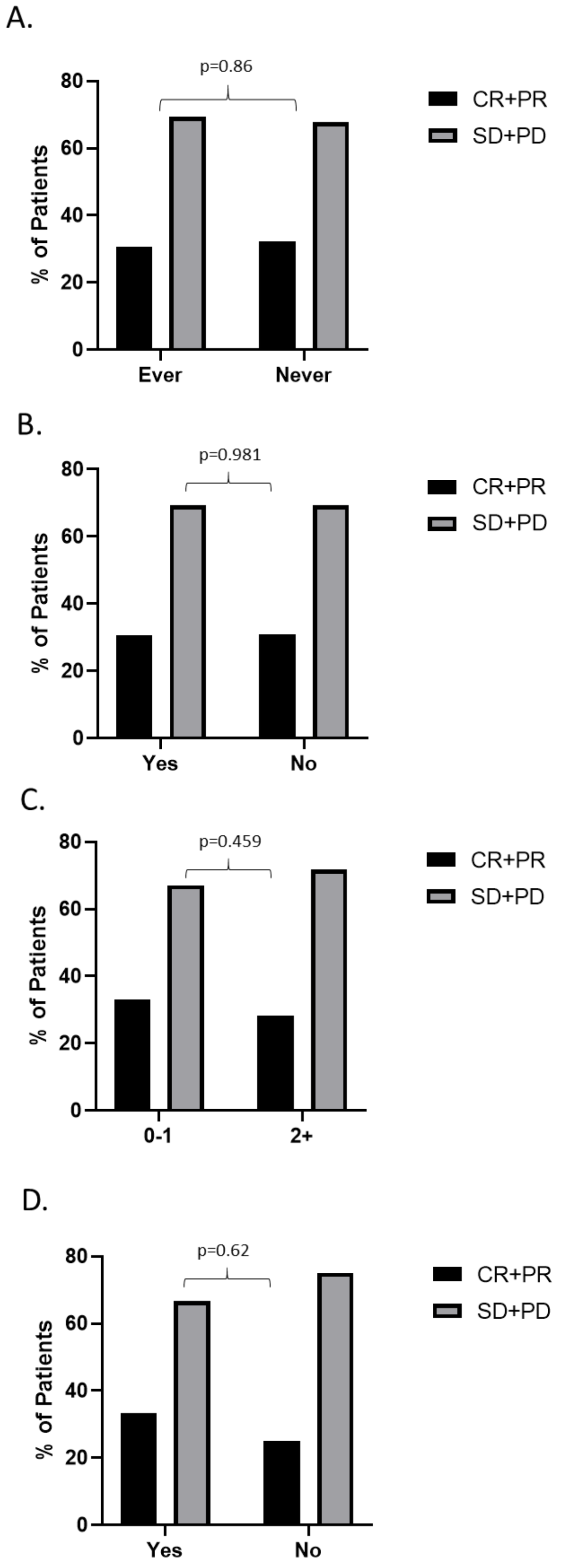


**Supplementary Figure 3. ORR as a function of cohort characteristics with significant differences among the race/ethnic groups for the entire cohort (ICI monotherapy and chemo/ICI combination therapy). A)** Smoking status, **B)** alcohol consumption, **C)** number of metastatic sites, and **D)** history of radiation treatment. CR- complete response; PR- partial response; SD- stable disease; PD- progressive disease.

**Supplementary Figure 4. ORR as a function of cohort characteristics with significant differences among the race/ethnic groups for the ICI monotherapy cohort. A)** Smoking status, **B)** alcohol consumption, **C)** number of metastatic sites, and **D)** history of radiation treatment. CR- complete response; PR- partial response; SD- stable disease; PD- progressive disease.

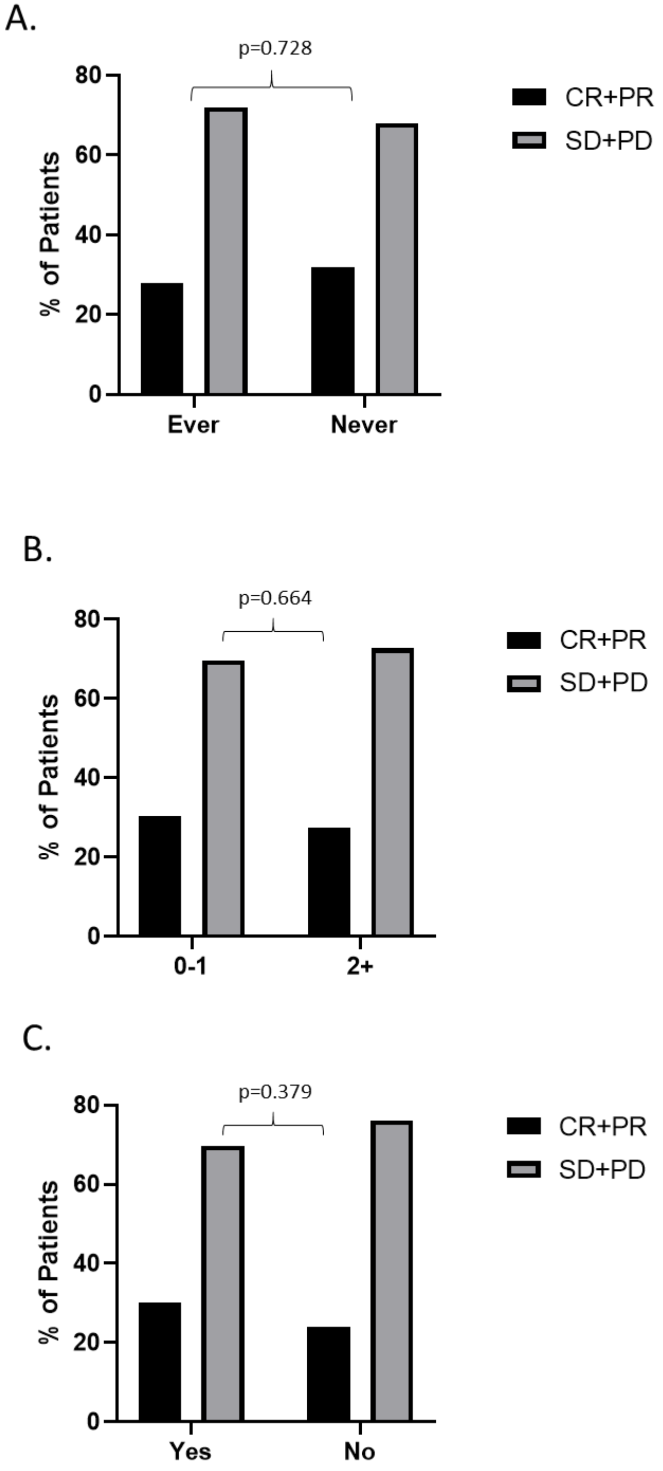
